# A Pilot Study of High-Intensity Interval Training in Older Adults with Treatment Naïve Chronic Lymphocytic Leukemia

**DOI:** 10.1101/2021.07.06.21259279

**Authors:** Grace MacDonald, Andrea Sitlinger, Michael A. Deal, Erik D. Hanson, Stephanie Ferraro, Carl F. Pieper, J. Brice Weinberg, Danielle M. Brander, David B. Bartlett

## Abstract

Chronic lymphocytic leukemia (CLL) is the most common leukemia in the USA, affecting predominantly older adults. CLL is characterized by low physical fitness, reduced immunity, and increased risk of secondary malignancies and infections. One approach to improving physical fitness and immune functions is participation in a structured exercise program. The aims of this pilot study were to determine the feasibility and outcomes of 12-weeks of high-intensity interval training (HIIT) combined with muscle endurance-based resistance training on older adults with treatment naïve CLL. We enrolled eighteen participants with CLL aged 64.9 (9.1) years and assigned them to groups depending on distance lived from our fitness center. Ten participants (4M/6F) completed HIIT and six participants (4M/2F) completed a non-exercising control group (Controls). HIIT consisted of three 30-minute treadmill sessions/week plus two concurrent 30-minute strength training sessions/week. We confirmed feasibility if >70% of HIIT participants completed >75% of prescribed sessions and prescribed minutes, and if >80% of high-intensity intervals were at a heart rate corresponding to 80% of aerobic capacity [139 (19) bpm]. Other outcomes included aerobic capacity, muscle strength and endurance, and natural killer (NK) cell recognition and killing of tumor cells. Results are presented as Hedge’s G effect sizes (*g*), with 0.2, 0.5 and 0.8 representing small, medium and large effects, respectively. Feasibility was achieved, with HIIT completing 5.0 (0.2) sessions/week and 99 (3.6)% of prescribed minutes/week at 142 (19)bpm. Following HIIT, leg strength (*g*=2.52), chest strength (*g*=1.15) and seated row strength (*g*=3.07) were 35.4%, 56.1% and 39.5% higher, respectively, while aerobic capacity was 3.8% lower (*g*=0.49) than changes for Controls. Similarly, following HIIT, *in vitro* NK-cell cytolytic activity against the K562 cell line (*g*=1.43), OSU-CLL cell line (*g*=0.95), and autologous B-cells (*g*=1.30) were 20.3%, 3.0% and 14.6% higher, respectively, than changes for Controls. We demonstrate that 12-weeks of HIIT combined with muscle endurance-based resistance training is feasible in older adults with untreated CLL and that HIIT has a large effect on muscle strength and important components of immune function.

## Introduction

With a median age at diagnosis of approximately 70 years, chronic lymphocytic leukemia (CLL) is the most prevalent adult leukemia in the USA [1-3]. The clinical presentation of CLL is diverse, and patients have a shorter life expectancy than age-matched healthy populations [4]. At present, there is no survival benefit from immediate or early therapy prior to established treatment indications, and most patients have a period of observation before therapy initiation [5, 6]. During the treatment naïve period, normal immune functions are impaired, increasing the risk of secondary malignancies and infections [7-9]. Furthermore, treatment naïve patients have low physical fitness that predicts poor survival following commencement of treatment [10].

One approach to concurrently improving physical fitness and immune functions is participation in a structured exercise program [11, 12]. Increasing physical activity levels and physical fitness are associated with improvements in therapy-related side effects, physical functioning, and quality of life for lymphoma patients [12-14]. Twelve weeks of moderate-intensity aerobic training improved physical function, fatigue, cardiovascular fitness, and QOL [14], while 36-weeks of exercise training was effective at improving balance and quality of life [13]. However, these studies included only 11.5% CLL patients [14] or 46% B-cell Non-Hodgkin’s Lymphoma patients, of which it is unclear how many had CLL [13]. No study that we are aware of has assessed the effects of exercise training on important immune functions for CLL patients.

In healthy older adults, higher physical activity and physical fitness is mostly associated with better immune functions [11, 15]. However, exercise-training interventions have more varied results, mainly because of the type, duration of and adherence to exercise. In clinical older adults, high-intensity interval training (HIIT) rapidly increases physical fitness, making it an attractive intervention for adults with low exercise capacity [16]. We, and others, have shown that HIIT in older patients with prediabetes or rheumatoid arthritis also reduces disease burden while improving important immune functions [16-20]. Therefore, HIIT may be an effective form of exercise to improve physical fitness and immune functions for CLL patients. Thus far, no study has assessed the effects of HIIT on physical fitness and immune functions in untreated CLL patients.

The aim of this study was to examine the feasibility of a 12-week HIIT intervention combined with muscle endurance-based resistance training in older adults with treatment naïve CLL. We used a HIIT intervention similar to one previously shown to provide immunomodulatory and aerobic fitness improvements [17, 18, 21] and a resistance program designed to improve muscular strength [22]. We hypothesized that >70% of participants assigned to HIIT would complete >75% (45 of 60 sessions) of prescribed sessions, and >75% (112.5 of 150 minutes/week) of prescribed weekly minutes. As exploratory aims, we sought to determine the effects of HIIT on aerobic capacity, skeletal muscle strength, and immune characteristics associated with resilience to CLL.

## Methods

This study was conducted as a two-arm, quasi-experimental pilot study at Duke University between August 2018 and February 2020. Figure 1 diagrams the study participant progression. All participants provided written informed consent prior to study testing, and the study was approved by the Duke University Medical Center Institutional Review Board. Participants were allocated to the exercise group (HIIT) or the control group (Controls), depending on the distance they lived from our facility used for the supervised exercise training. If participants lived >35 miles away, they were assigned to the control arm as this was deemed too far to travel three times per week.

**Figure 1.**
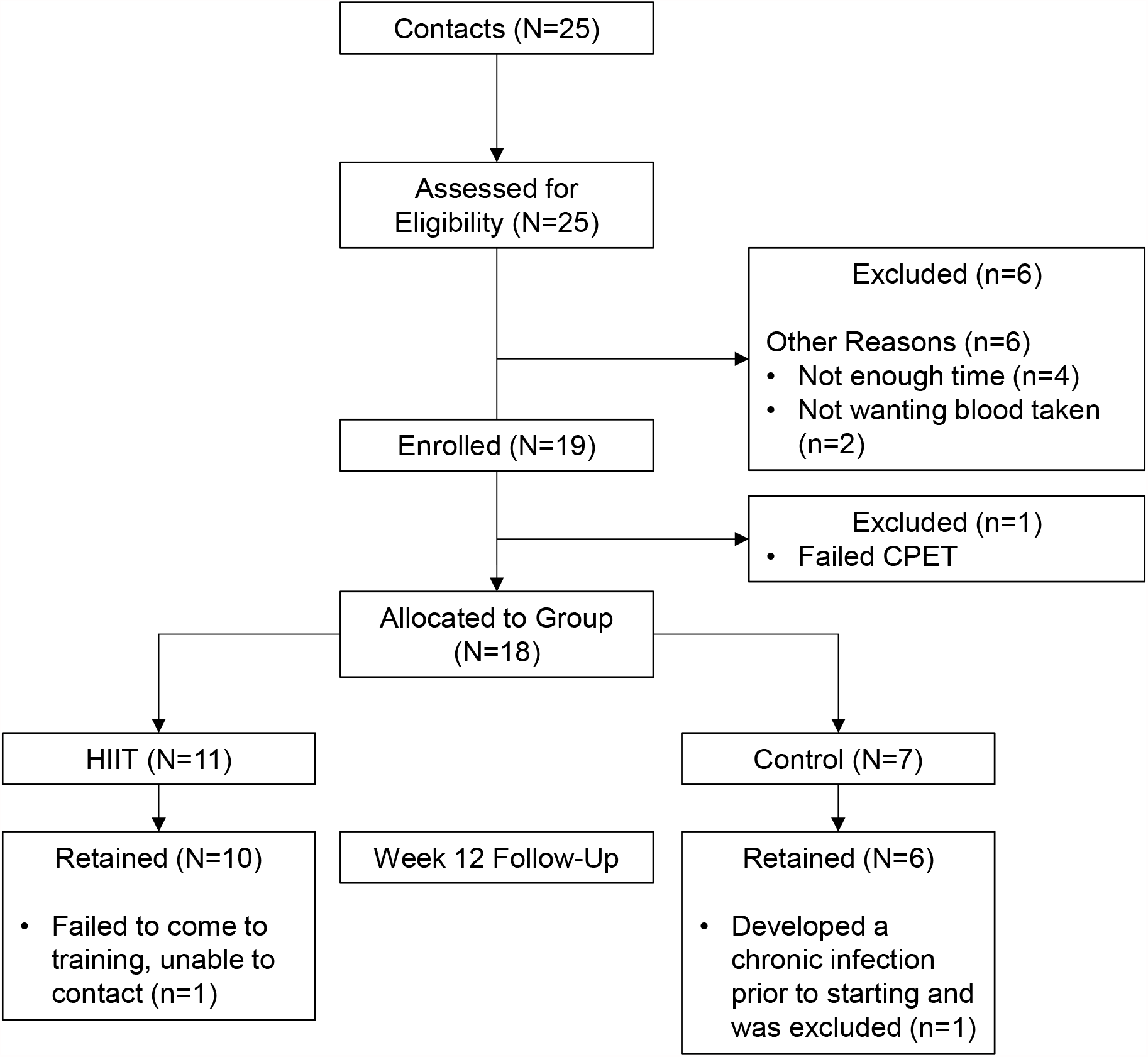
Consolidated Standards of Reporting Trials (Consort) Diagram

### Eligibility

Eligibility requirements included (1) no abnormal cardiac findings observed during a maximal cardiopulmonary exercise test (CPET) that would prevent participants from exercising safely at high exercise intensities (see below for more details); (2) male or female with confirmed diagnosis of CLL as per the International Workshop on CLL Guidelines [23]; (3) no history of prior treatment of CLL; (4) able to walk on a treadmill; (5) no clinical evidence of significant disease progression with consideration for first line therapy within 6 months; (6) no systemic glucocorticoid therapy within the past 7 days; (7) no malignancy diagnosed within 3 years of study enrollment except for adequately treated basal, squamous cell carcinoma or non-melanomatous skin cancer, carcinoma in situ of the cervix, superficial bladder cancer not treated with intravesical chemotherapy or BCG within 6 months, localized prostate cancer; (8) no absolute contra-indications to exercise, including recent (<6 months) unstable angina, uncontrolled dysrhythmias causing symptoms or hemodynamic compromise, symptomatic aortic stenosis, uncontrolled symptomatic heart failure, acute pulmonary embolus, acute myocarditis or pericarditis, suspected or known dissecting aortic aneurism, or acute systemic infection; (9) no orthopedic limitations, musculoskeletal disease and/or injury. Due to the nature of the study, participants with known joint, muscle or other orthopedic limitations that restrict physical activity were excluded; (10) no diabetes mellitus or chronic obstructive pulmonary disease; and (11) no uncontrolled hypertension (blood pressure ≥160/90).

### Fitness, Strength, and Body Composition

#### Cardiorespiratory Fitness

We used a medically supervised cardiopulmonary exercise test (CPET) to assess cardiorespiratory fitness and cardiac health, including continuous 12-lead electrocardiogram assessment and breath-by-breath metabolic analyses (ParvoMedics, UT, USA) as previously described [24]. Aerobic capacity (VO_2peak_) was determined by a graded maximal treadmill test starting at 2 mph/0% grade with increasing speed and/or grade such that the metabolic demand increased at approximately 3.5 mL/kg/min until volitional exhaustion. We confirmed a valid test by either a respiratory exchange ratio (RER) of >1.1 [mean RER: 1.2 (0.1)] or a rating of perceived exertion of ≥ 17 [RPE: mean 18 (1)]. We recorded blood pressure before, after the CPET, and at the end of each stage during the CPET. **Body Composition.** Total body mass, fat mass and lean mass were estimated by air-displacement plethysmography (BodPod System; Life Measurement Corporation, Concord, CA) [24]. **Muscle Strength.** We determined estimated maximal muscle strength and muscular endurance using three machine-weights – leg press, chest press, and seated row. At the initial visit, each exercise was explained to the participant and demonstrated by the exercise physiologist. Briefly, a warm-up set of 8-10 repetitions at 40-50% of predicted 1RM [25], before a priming set of 5-7 reps at 50-60% 1RM was performed. Participants were then instructed to perform the maximum number of repetitions at 80-85% 1RM. If after the first attempt over 8 repetitions were performed, the weight was increased for the next set until no more than 8 repetitions could be completed. Participants were given 3-5 minutes rest in between each set. Predicted 1 RM was calculated and the participant performed as many repetitions as possible (muscular endurance) at 70% of 1 RM on each machine [26]. **Physical Activity Levels.** We assessed participant’s habitual physical activity exposure by 7-day continuous wear of a wrist-based accelerometer (Garmin Vivosmart 3, Garmin, Kansas, USA) prior to and following the intervention. We assessed total steps/day during waking hours each day as a measure of habitual physical activity. Sleep time was determined by the device and confirmed by a sleep diary provided by each participant. All of the above tests and the blood samples were completed before training and 24 hours after the participants last exercise session.

### 12-Week Exercise Training Program

The control group were informed to continue their daily activities and not participate in structured exercise programs. Participants assigned to the exercise group completed supervised training 3 times per week for 12-weeks. Two of the sessions were approximately 1 hour, with 30 minutes for HIIT and 30 minutes for resistance training, and 1 session approximately 30 minutes for HIIT only (**Table 1**). Exercise intensities were determined from the CPET using VO_2_ reserve calculated as previously described [27]. Participants were given between 3 and 6 sessions to become accustomed to the exercise (30-to 45-second intervals at target heart rates, total time 30 minutes). Each exercise bout consisted of a 5-minute warm-up and 5-minute cool down as part of the total session. Intervals were designed to elicit a heart rate corresponding to 80-90% of VO_2_ reserve (high intensity intervals) and 50-60% VO_2_ reserve (active recovery). Heart rate was recorded continuously during each session (Polar OH1, Polar, USA). Speeds did not exceed walking pace (1 – 4.8 mph), and if heart rate was not achieved by the end of the interval, the gradient (1 – 15%) was elevated to increase heart rate. High intensity intervals were between 60 and 90 seconds followed by active recovery intervals of a similar duration. Rather than controlling each session for energy expenditure, we adjusted total intervals per session so that the aerobic exercise duration each session was 30 minutes. Ratings of perceived exertion were detailed at the end of each high intensity interval. Resistance training exercises targeted major muscle groups in the upper and lower body using the leg press, chest press, and seated row machines. Following a warm-up of 10 repetitions at 40-60% 1 RM, sets consisted of as many repetitions possible at 70% 1 RM were completed. Once participants were able to perform 20 repetitions or more, the weight was increased for the next session by 2-5 kg. We adopted a flexible scheduling protocol to ensure participants could complete all 3 prescribed sessions/week (6am to 6pm, any day of 5 days per week or per request of the participant), and supervision by an exercise physiologist. If a participant was unable to complete the entire session, the number of minutes completed in that session was documented.

**Table 1.**
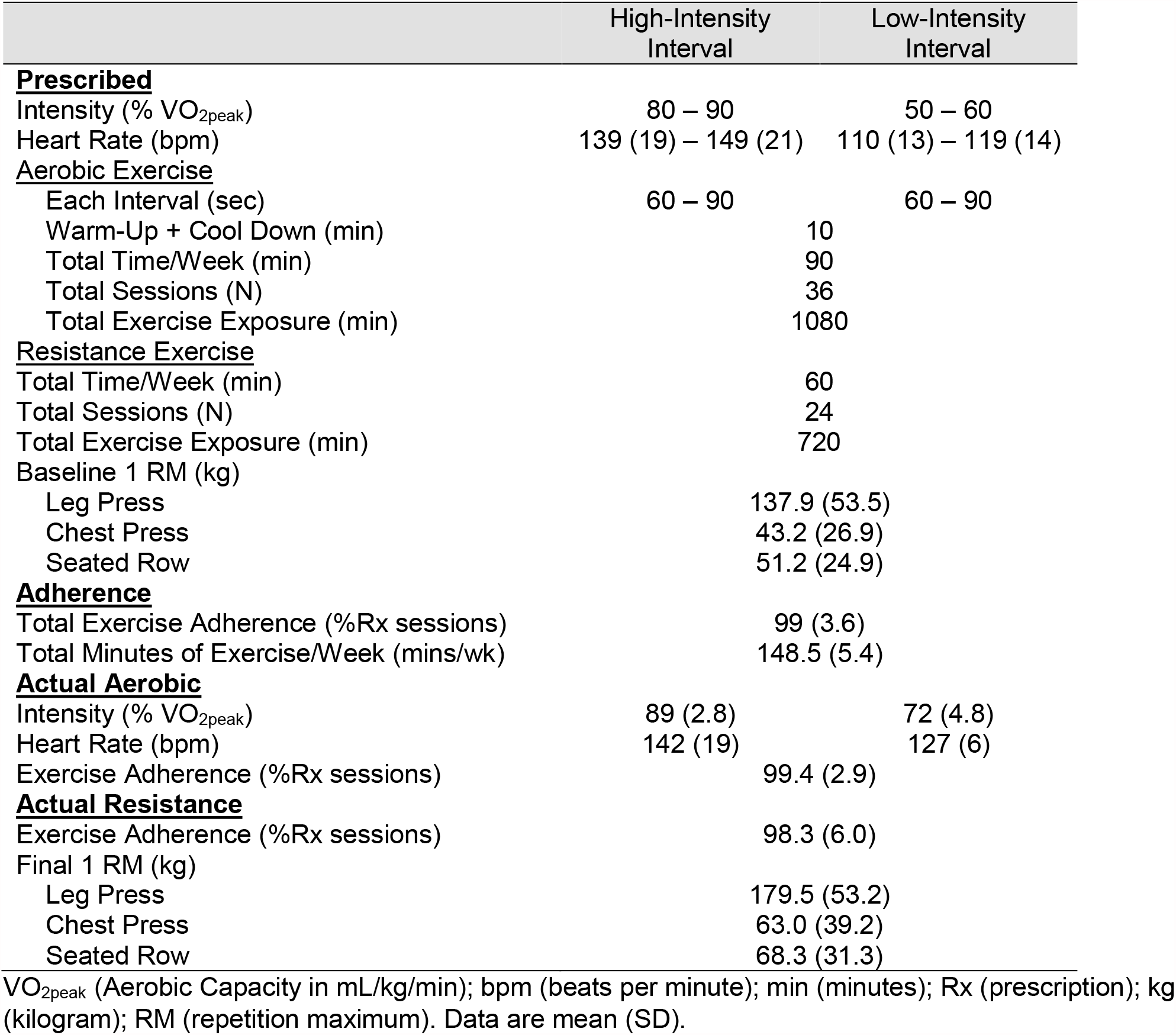
Exercise Prescription and results of the intervention

### Feasibility, Fidelity, Compliance, and Safety

We defined feasibility by participant adherence. Specifically, feasibility was achieved if >70% of participants assigned to HIIT could complete >75% (45 of 60 sessions) of prescribed sessions and complete >75% (112.5 of 150 minutes/week) of prescribed weekly minutes. Before and after every session, adverse events were monitored by determining if participants had any injuries, nausea, infections, pain, and shortness of breath. We confirmed fidelity if >80% of all prescribed testing was completed. Fidelity was calculated as the percentage of all tests each participant completed at baseline, and at the follow-up after the total number of required tests were completed (e.g. aerobic fitness, muscle strength, body composition, blood samples). We confirmed compliance if >80% of all high-intensity intervals were completed at a heart rate corresponding to 80% VO_2peak_. Each high-intensity interval average heart was recorded and compared against prescribed heart rates determined from aerobic fitness testing.

### Blood Samples

Participants arrived for blood draws at the same time before and after the intervention having consumed the same diet the day prior to the visit. They were instructed to not eat or drink anything besides water the morning of their visit. A total of 50 mL blood was taken for immediate processing. Complete blood differential counts were measured in EDTA anticoagulated whole blood using a clinical hematology analyzer (Sysmex, USA).

### Immune Cell Isolation

Peripheral blood mononuclear cells (PBMCs) were isolated within 1 hour of collection, from heparin anticoagulated blood following density gradient centrifugation, as previously described [28]. Briefly, blood was diluted 1:1 with sterile phosphate buffer saline (PBS) before being layered on Ficoll-Paque Plus and processed according to manufacturer guidelines (Sigma-Aldrich, MO, USA). PBMC viability was determined by trypan blue exclusion and was consistently >98%. A sample of PBMCs were processed for cryopreservation using standard methods (fetal bovine serum (FBS) + 10% DMSO) and stored in liquid nitrogen.

We isolated CD19+ B-cells from 5mL of blood using negative selection density gradient centrifugation using the manufacturer’s guidelines (RosetteSep Human B Cell Enrichment Kit, STEMCELL technologies, MA, USA). Briefly, blood was incubated for 20 minutes with a tetrameric antibody complex that recognizes non-B-cells and glycophorin A on RBCs. Blood was diluted with sterile PBS and layered on Ficoll-Paque. Following centrifugation, only the negatively enriched B-cells at the interphase were removed, washed and analyzed for purity using CD19+ flow cytometry staining and viability by trypan exclusion. Purity and viability were consistently >98%. A sample of B-cells was processed for cryopreservation, with the remaining cells used for the target population in the NK-cell cytotoxicity assays (see below). These cells were maintained at 1 × 10^6^ cells/mL in complete media (RPMI 1640 + 2mM L-glutamine, 100 U/mL penicillin, 100 µg/mL streptomycin, with 10% FBS).

### Measurement of PBMC Subsets by Flow Cytometry

We determined frequencies of PBMC subset (T-cells, B-cells, CLL-cells, NK-cells and monocytes) using a 3-laser BD FACSCanto II (BD Bioscience, USA) flow cytometer. Flow cytometry was completed at the Duke Cancer Institute Core Facility, which maintained daily quality controls of the machine. We analyzed 10,000 cells of interest and assessed the data with FCS Express 6 (FCS Express, USA). Lymphocytes and monocytes were gated using standard forward versus side scatter properties. We identified NK-cells as CD3^neg^/CD56+, T-cells as either CD3+/CD4+ or CD3+/CD8+, B-cells as CD3^neg^/CD19+/CD5^neg^, CLL-cells as CD3^neg^/CD19+/CD5+, and monocytes as CD14+ cells. We further classified monocytes as classic (CD14+/CD16^neg^), intermediate (CD14+/CD16+) or non-classic (CD14+/CD16++). PBMCs (1 × 10^6^ cells/mL in PBS/1% BSA) were incubated on ice for 30 minutes with combinations of the following fluorochrome conjugated monoclonal antibodies, or relevant isotype concentration-matched isotype controls. All antibodies were titrated prior to assessing samples and single color and flow minus one (FMOs) tubes were used for compensation. We used 0.75µg/mL CD3 Pacific Blue (Clone UCHT1; BD Bioscience, NC, USA), 0.2µg/mL CD56-PE (Clone 5.1H11, BioLegend, CA, USA), 1µg/mL CD4-PE (Clone OKT4; BioLegend), 10µg/mL CD8 FITC (Clone OKT8; Thermofisher, MA, USA), 1.5µg/mL CD19-APC-Cy7 (Clone HIB19; BioLegend), 3µg/mL CD5-APC (Clone UCHT2; BioLegend), 1µg/mL CD14-PcB (Clone TuK4; Thermofisher), and CD16-FITC (Clone 3G8, BD Bioscience). After incubations, we washed samples once and resuspended them in 300µL of 4% formaldehyde for 20 minutes, before flow cytometry.

### NK-Cell Cytolytic Activity

We assessed NK-cell cytotoxicity (NKCC) by three-color flow cytometry using adapted methods of Hazeldine *et al*. [29]. Briefly, we isolated NK-cells from PBMC samples by negative magnetic selection using MACS® technology (Human NK Cell Isolation Kit, Miltenyi Biotec, MD, USA), and resuspended them in complete media at 1 × 10^6^ cells/mL. NK-cells were assessed for purity via CD3^neg^/CD56^+^ flow cytometry staining and viability by trypan exclusion. Purity and viability were consistently >95%. Target cells were the MHC-I deficient cell line K562 (ATCC, VA, USA), the EBV transformed stable CLL-like cell line OSU-CLL, from the Byrd Lab at Ohio State University [30], and autologous B-cells isolated from PBMC samples (see above). K562 and OSU-CLL cells were maintained in complete media at 37° C in a humidified 5% CO_2_ incubator. We stained K562 and OSU-CLL cells with anti-CD71-PE-Cy7, while autologous B-cells were stained with anti-CD19-APC-Cy7 and anti-CD5-APC, prior to the assay beginning, and similar to previous methods [31]. K562 and OSU-CLL cell passage was identical for pre and post-intervention. NK-cells and target cells were co-cultured at effector: target (E:T) ratios of 10:1, 5:1, and 0:1 (spontaneous death) for 4 hours at 37°C/5% CO_2_. After incubation, we pelleted cells and resuspended them in PBS/1% BSA containing anti-CD56-PE for 20 minutes on ice. Cells were then washed and resuspended in PBS, and stained for 5 minutes with 125 nM sytox® blue dead cell stain (ThermoFisher, MA, USA) before immediate analyses by flow cytometry. NKCC was determined by the number of lysed target cells (defined as sytox blue positive) within a population of 5000 cells, with percentage-specific lysis calculated as the number of lysed target cells (TL) minus lysed target cells without effector cells (SL), divided by the number of cells recorded (C#) and multiplied by 100 ((TL-SL/C#) x 100) [29].

### Disease Characteristics

We obtained clinical indices from patients’ medical records. Indices included disease duration, cytogenetics, IGHV mutation status, and CD38 expression. The CLL-IPI score, calculated as previously described [32]. We prepared plasma from blood immediately after venesection, and samples were stored at minus 80°C until analyzed. All plasma analyses were completed within the Biomarker Core Facilities of the Duke Molecular Physiology Institute. Concentrations of soluble CD20 (sCD20: pg/mL) and intercellular adhesion molecule 1 (ICAM-1: ng/mL) were determined in duplicate using a human sandwich immunoassay according to the manufacturer’s instructions (Meso Scale Discovery, Rockville, MD). β2-microglobulin (B2M: µg/mL) was determined in duplicate using a commercially available ELISA (R&D Systems, Minneapolis, MN). The lower limits of detection (LLOD) were sCD20 (30.5 pg/mL), ICAM-1 (2.60 ng/mL), and B2M (0.132 µg/mL). All samples had concentrations greater than the LLOD with the exception of sCD20, for which 81% of samples were above the LLOD.

### Statistics

This pilot, single-center study in which participants were not randomly assigned was quasi-experimental. All analyses were conducted using IBM SPSS version 23.0 (Armonk, NY, United States). Normality was assessed using Kolmogorov-Smirnov analysis. For variables violating normality, natural log transformation was performed or non-parametric analyses was employed. Descriptive statistics are shown for baseline characteristics and compared across groups using independent T-tests (or Wilcoxon sum of ranks) for continuous variables and goodness-of-fit chi-square for categorical. For each participant, adherence was calculated as 1) the number of sessions attended/number of prescribed sessions and multiplied by 100 and 2) the average number of weekly exercise minutes/number of prescribed weekly minutes and multiplied by 100. Differences in mean percent change by group in fitness and immune function was assessed by ANCOVA, where the baseline value of each dependent variable was included as a covariate. Because of the risk of recruitment bias, we have not reported *p*-values throughout. Instead, statistics are presented as mean (standard deviation (SD)) or mean differences and 95% confidence intervals, unless otherwise indicated, and presented with effect sizes. Effect sizes were calculated as Hedges’ *G* (*g*) for comparisons between baseline characteristics, and mean intervention differences [33, 34]. We considered small, medium and large effect sizes if *g* equated to 0.2, 0.5 and 0.8, respectively.

## Results

### Participants

Baseline group demographics, disease related blood markers and disease characteristics are presented in Table 2. There were sixteen participants [8 men and 8 women, mean age of 64.9 (9.1) years] with stable, confirmed treatment-naïve CLL who completed the 12-week study. Mean years since diagnosis was 6.3 years (range: 0.5 – 24 years). Participants were mostly Rai stage 0 or 1 (81.2%), and those with CLL-IPI scores were mostly 0 or 1. One participant in the control group had a CLL-IPI score of 3 and one in the HIIT group a score of 2. Fourteen (87.5%) participants had favorable cytogenetics as defined as either normal, del13q, monosomy 13, trisomy 12 and/or del11q. *IGHV* was assessed in ten participants (62.5%) and *TP53* was assessed in six participants (37.5%). Groups were similar at baseline for physical fitness and most immune cell characteristics (**Table 3**). Large effect sizes were observed between groups for absolute monocyte counts, which were 37.5% lower in the HIIT group (95% CI: [-0.6, -0.01], *g*=1.08), and the ratio of T-cells to monocytes which were 40.7% higher in the HIIT group (95% CI: [-0.9, 3.1], *g*=0.82). Large effect sizes were observed between groups for the frequency (% of lymphocytes) of CD8+ T-cells, which were 6.8% lower in the HIIT group (95% CI: [-15.2, 1.6], *g*=0.92) and the frequency of intermediate monocytes, which were 1.6% lower in the HIIT group (95% CI: [-3.7, 0.5], *g*=0.82). Absolute counts of NK-cells were 40.1% lower in the HIIT group (95% CI: [-0.5, 0.1], *g*=0.89), while NK-cell specific lysis of autologous B-cells was 11.4% lower in the HIIT group (95% CI: [-1.4, 21.5], *g*=1.22).

**Table 2.**
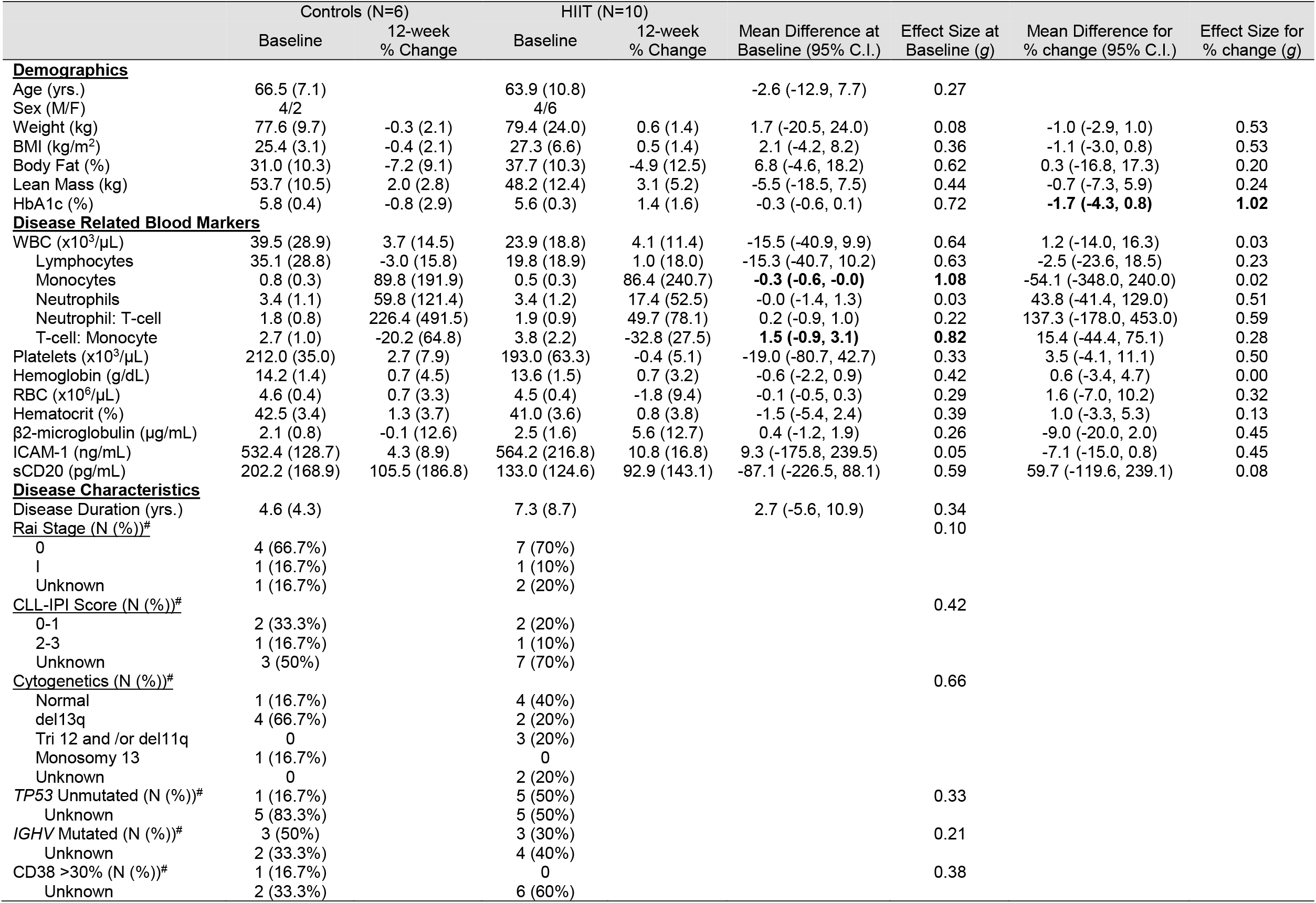
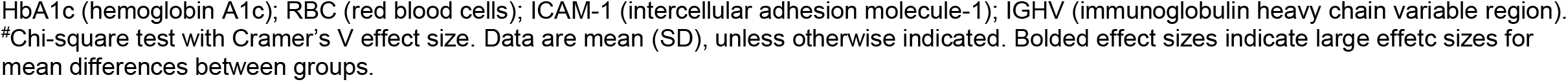
Baseline and percentage changes upon study completion for demographics and clinical characteristics of participants completing the intervention.

|  | Controls (N=6) |  | HIIT (N=10) |  | Mean Difference at Baseline (95% C.I.) | Effect Size at Baseline (g) | Mean Difference for % change (95% C.I.) | Effect Size for % change (g) |
| --- | --- | --- | --- | --- | --- | --- | --- | --- |
|  | Baseline | 12-week % Change | Baseline | 12-week % Change |  |  |  |  |
| **Demographics** |  |  |  |  |  |  |  |  |
| Age (yrs.) | 66.5 (7.1) |  | 63.9 (10.8) |  | -2.6 (-12.9, 7.7) | 0.27 |  |  |
| Sex (M/F) | 4/2 |  | 4/6 |  |  |  |  |  |
| Weight (kg) | 77.6 (9.7) | -0.3 (2.1) | 79.4 (24.0) | 0.6 (1.4) | 1.7 (-20.5, 24.0) | 0.08 | -1.0 (-2.9, 1.0) | 0.53 |
| BMI (kg/m<sup>2</sup>) | 25.4 (3.1) | -0.4 (2.1) | 27.3 (6.6) | 0.5 (1.4) | 2.1 (-4.2, 8.2) | 0.36 | -1.1 (-3.0, 0.8) | 0.53 |
| Body Fat (%) | 31.0 (10.3) | -7.2 (9.1) | 37.7 (10.3) | -4.9 (12.5) | 6.8 (-4.6, 18.2) | 0.62 | 0.3 (-16.8, 17.3) | 0.20 |
| Lean Mass (kg) | 53.7 (10.5) | 2.0 (2.8) | 48.2 (12.4) | 3.1 (5.2) | -5.5 (-18.5, 7.5) | 0.44 | -0.7 (-7.3, 5.9) | 0.24 |
| HbA1c (%) | 5.8 (0.4) | -0.8 (2.9) | 5.6 (0.3) | 1.4 (1.6) | -0.3 (-0.6, 0.1) | 0.72 | **-1.7 (-4.3, 0.8)** | **1.02** |
| **Disease Related Blood Markers** |  |  |  |  |  |  |  |  |
| WBC (x10<sup>3</sup>/μL) | 39.5 (28.9) | 3.7 (14.5) | 23.9 (18.8) | 4.1 (11.4) | -15.5 (-40.9, 9.9) | 0.64 | 1.2 (-14.0, 16.3) | 0.03 |
| Lymphocytes | 35.1 (28.8) | -3.0 (15.8) | 19.8 (18.9) | 1.0 (18.0) | -15.3 (-40.7, 10.2) | 0.63 | -2.5 (-23.6, 18.5) | 0.23 |
| Monocytes | 0.8 (0.3) | 89.8 (191.9) | 0.5 (0.3) | 86.4 (240.7) | **-0.3 (-0.6, -0.0)** | **1.08** | -54.1 (-348.0, 240.0) | 0.02 |
| Neutrophils | 3.4 (1.1) | 59.8 (121.4) | 3.4 (1.2) | 17.4 (52.5) | -0.0 (-1.4, 1.3) | 0.03 | 43.8 (-41.4, 129.0) | 0.51 |
| Neutrophil: T-cell | 1.8 (0.8) | 226.4 (491.5) | 1.9 (0.9) | 49.7 (78.1) | 0.2 (-0.9, 1.0) | 0.22 | 137.3 (-178.0, 453.0) | 0.59 |
| T-cell: Monocyte | 2.7 (1.0) | -20.2 (64.8) | 3.8 (2.2) | -32.8 (27.5) | **1.5 (-0.9, 3.1)** | **0.82** | 15.4 (-44.4, 75.1) | 0.28 |
| Platelets (x10<sup>3</sup>/μL) | 212.0 (35.0) | 2.7 (7.9) | 193.0 (63.3) | -0.4 (5.1) | -19.0 (-80.7, 42.7) | 0.33 | 3.5 (-4.1, 11.1) | 0.50 |
| Hemoglobin (g/dL) | 14.2 (1.4) | 0.7 (4.5) | 13.6 (1.5) | 0.7 (3.2) | -0.6 (-2.2, 0.9) | 0.42 | 0.6 (-3.4, 4.7) | 0.00 |
| RBC (x10<sup>6</sup>/μL) | 4.6 (0.4) | 0.7 (3.3) | 4.5 (0.4) | -1.8 (9.4) | -0.1 (-0.5, 0.3) | 0.29 | 1.6 (-7.0, 10.2) | 0.32 |
| Hematocrit (%) | 42.5 (3.4) | 1.3 (3.7) | 41.0 (3.6) | 0.8 (3.8) | -1.5 (-5.4, 2.4) | 0.39 | 1.0 (-3.3, 5.3) | 0.13 |
| β2-microglobulin (μg/mL) | 2.1 (0.8) | -0.1 (12.6) | 2.5 (1.6) | 5.6 (12.7) | 0.4 (-1.2, 1.9) | 0.26 | -9.0 (-20.0, 2.0) | 0.45 |
| ICAM-1 (ng/mL) | 532.4 (128.7) | 4.3 (8.9) | 564.2 (216.8) | 10.8 (16.8) | 9.3 (-175.8, 239.5) | 0.05 | -7.1 (-15.0, 0.8) | 0.45 |
| sCD20 (pg/mL) | 202.2 (168.9) | 105.5 (186.8) | 133.0 (124.6) | 92.9 (143.1) | -87.1 (-226.5, 88.1) | 0.59 | 59.7 (-119.6, 239.1) | 0.08 |
| **Disease Characteristics** |  |  |  |  |  |  |  |  |
| Disease Duration (yrs.) | 4.6 (4.3) |  | 7.3 (8.7) |  | 2.7 (-5.6, 10.9) | 0.34 |  |  |
| Rai Stage (N (%))<sup>#</sup> |  |  |  |  |  | 0.10 |  |  |
| 0 | 4 (66.7%) |  | 7 (70%) |  |  |  |  |  |
| I | 1 (16.7%) |  | 1 (10%) |  |  |  |  |  |
| Unknown | 1 (16.7%) |  | 2 (20%) |  |  |  |  |  |
| CLL-IPI Score (N (%))<sup>#</sup> |  |  |  |  |  | 0.42 |  |  |
| 0-1 | 2 (33.3%) |  | 2 (20%) |  |  |  |  |  |
| 2-3 | 1 (16.7%) |  | 1 (10%) |  |  |  |  |  |
| Unknown | 3 (50%) |  | 7 (70%) |  |  |  |  |  |
| Cytogenetics (N (%))<sup>#</sup> |  |  |  |  |  | 0.66 |  |  |
| Normal | 1 (16.7%) |  | 4 (40%) |  |  |  |  |  |
| del13q | 4 (66.7%) |  | 2 (20%) |  |  |  |  |  |
| Tri 12 and /or del11q | 0 |  | 3 (20%) |  |  |  |  |  |
| Monosomy 13 | 1 (16.7%) |  | 0 |  |  |  |  |  |
| Unknown | 0 |  | 2 (20%) |  |  |  |  |  |
| TP53 Unmutated (N (%))<sup>#</sup> | 1 (16.7%) |  | 5 (50%) |  |  | 0.33 |  |  |
| Unknown | 5 (83.3%) |  | 5 (50%) |  |  |  |  |  |
| IGHV Mutated (N (%))<sup>#</sup> | 3 (50%) |  | 3 (30%) |  |  | 0.21 |  |  |
| Unknown | 2 (33.3%) |  | 4 (40%) |  |  |  |  |  |
| CD38 >30% (N (%))<sup>#</sup> | 1 (16.7%) |  | 0 |  |  | 0.38 |  |  |
| Unknown | 2 (33.3%) |  | 6 (60%) |  |  |  |  |  |

HbA1c (hemoglobin A1c); RBC (red blood cells); ICAM-1 (intercellular adhesion molecule-1); IGHV (immunoglobulin heavy chain variable region).

**Table 3.**
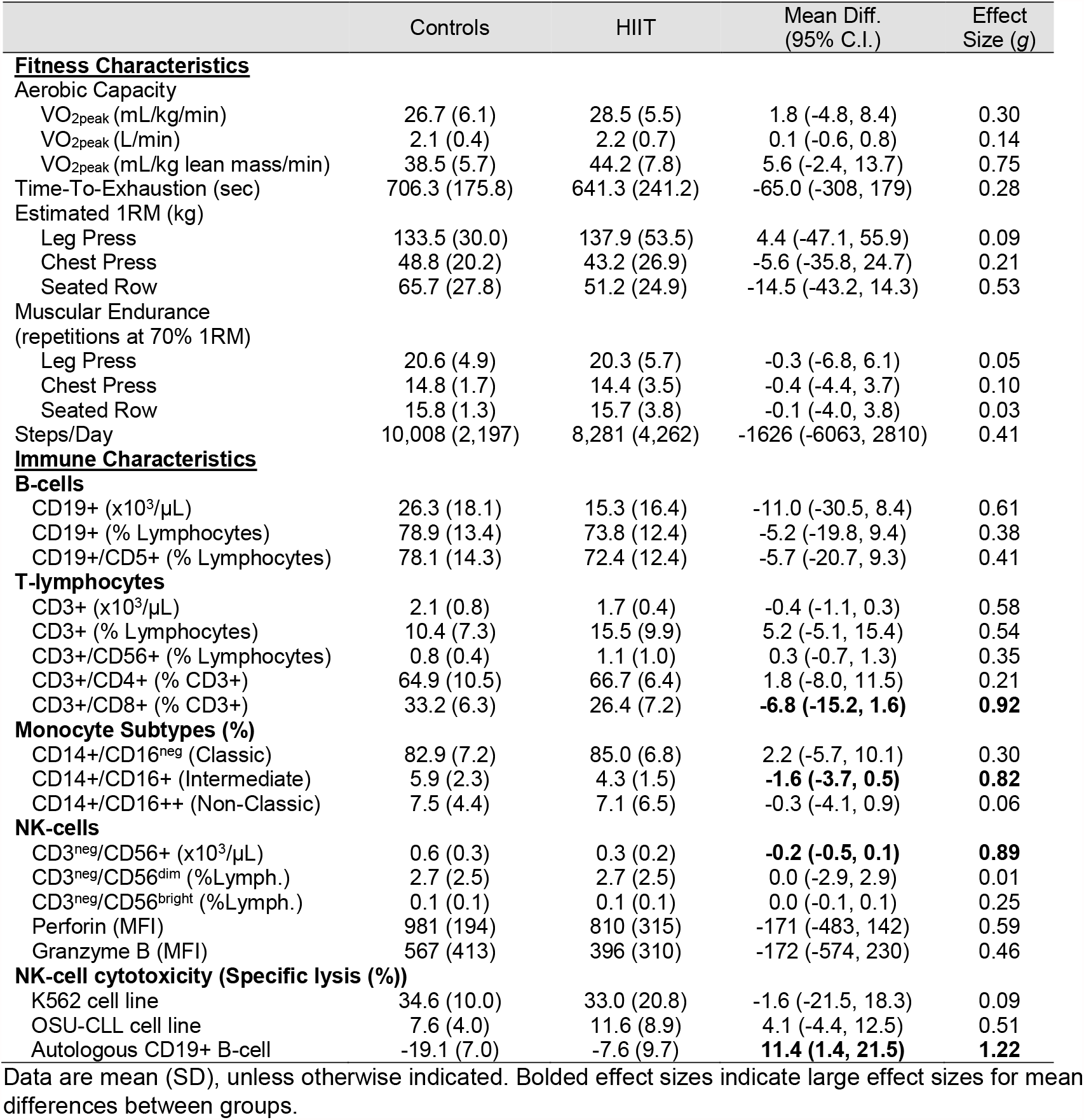
Baseline fitness and immune characteristics of participants completing the intervention.

### Accrual and Retention

Recruitment rate was 76% over the 18-month period (Figure 1). A total of six screened participants declined participation because they did not have enough time for participation (n=4) or did not want additional blood taken (n=2). Nineteen participants were consented into the study. One participant had an abnormal CPET and was referred to cardiology before being excluded from the study. Of the 18 participants assigned to the study groups (11 to HIIT, 7 to Control), 16 completed the study. One Control participant developed a chronic skin infection (hives) shortly after the CPET, but before training begun, and was excluded. One participant in the HIIT group failed to come to training or to respond to repeated attempts to be contacted and was excluded. The remaining 16 participants completed the 12-week program.

### Feasibility, Fidelity, Compliance, and Safety

Ten HIIT participants completed the study and all fulfilled feasibility criteria, with 5.0 (0.2) exercise sessions/week completed [99 (3.6)%]. This consisted of 3.0 (0.2) treadmill sessions completed and 2.0 (0.1) resistance sessions completed. Participants completed 148.5 (5.4) exercise minutes/week completed or 99 (3.6)% of prescription, with 100% of the participants completing greater than 75% of prescribed minutes. Our study fulfilled participant fidelity, with 95.0 (7.1)% and 86.3 (7.1)% of all required tests completed at baseline and at 12-weeks, respectively.

We determined program safety by recording the incidence and severity of pain or injuries throughout the program. There were no adverse events recorded during any of the exercise sessions. At the beginning of the study, 100% of HIIT participants reported minor muscle soreness due to the resistance and aerobic exercise but were considered normal reactions to exercise training. Incidences that required brief rescheduling of training sessions included edema in the arm (N=1), knee pain (N=1), upper respiratory infection (N=1), groin tenderness (N=1), and mild foot pain (N=1). These participants reduced training load until the situation resolved (all <1 week). One participant was dizzy and nauseous >3 hours following the baseline CPET and was admitted to the emergency department (ED). Following consultation with ED and oncology, it was deemed that the CPET was not the cause of the symptoms, and the participant was allowed to continue in the study after discharge from the ED.

Compliance was achieved, with 100% of participants completing >80% of high-intensity intervals at the prescribed heart rate. Mean high-intensity heart rate was 142 (19)bpm compared to the prescription of 139 (19)bpm at 80% VO_2peak_ and 149 (21)bpm at 90% VO_2peak_.

### Effects of HIIT

Upon completing the 12-weeks, we observed similar percent changes between Controls and HIIT for disease related blood markers and most demographics, with HIIT having a large effect on HbA1c that was 1.7% higher than Controls (Table 2: 95% CI = [-4.3, 0.8], *g*=1.02).

#### Aerobic Capacity

HIIT had a small-medium effect on aerobic capacity. As compared to Controls, the percentage change for relative aerobic capacity was 3.8% lower following HIIT (Fig. 2A: 95% CI = [-15.8, 8.1], *g*=0.49), while absolute aerobic capacity was 3.5% lower following HIIT (Fig. 2B: 95% CI = [-14.3, 7.3], *g*=0.43), and lean mass adjusted relative aerobic capacity was 4.0% lower following HIIT (Fig. 2C: 95% CI = [-20.1, 12.1], *g*=0.49).

**Figure 2.**
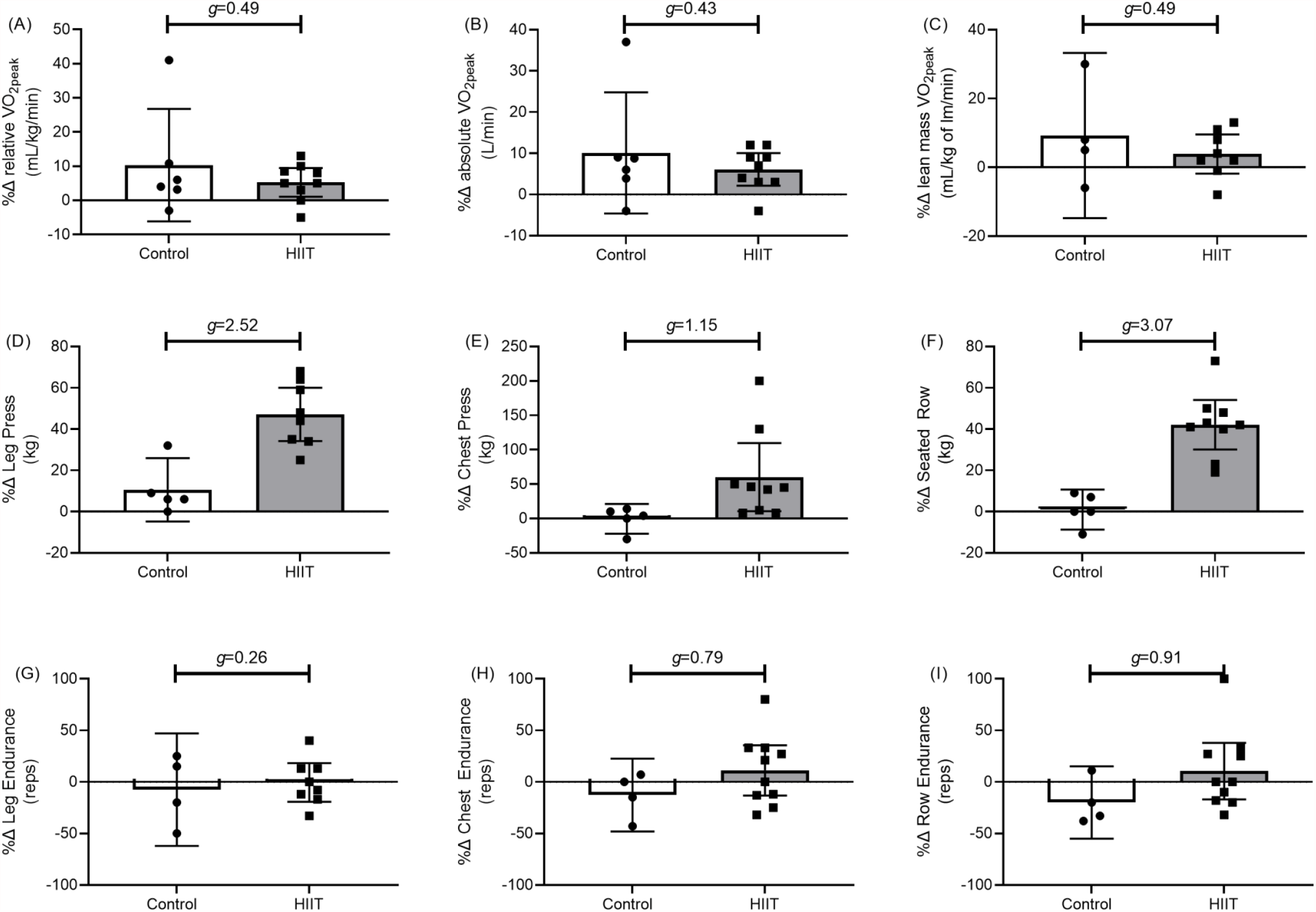
Mean (95% C.I.) percentage change (%Δ) with Hedges G (*g*) group differences between HIIT and Controls for cardiorespiratory fitness (A-C), estimated 1 repetition maximum strength (D-F), and number of repetitions completed at 70% of 1 RM (G-I).

#### Maximum Strength

HIIT had a large effect on muscle strength. As compared to Controls, the percentage change for maximal leg strength was 35.4% higher following HIIT (Fig. 2D: 95% CI = [17.3, 53.5], *g*=2.52), while maximal chest strength was 56.1% higher following HIIT (Fig. 2E: 95% CI = [-8.1, 120.3], *g*=1.15), and maximal seated row strength was 39.5% higher following HIIT (Fig. 2F: 95% CI = [22.6, 56.5], *g*=3.07).

#### Muscular Endurance

HIIT had a medium-large effect on upper body muscular endurance. As participants in the HIIT group increased muscular strength (Table 1), the relative weight used to test muscular endurance increased also. As compared to Controls, the percentage change for leg muscular endurance was 10.4% higher following HIIT (Fig. 2G: 95% CI = [-27.3, 48.2], *g*=0.26), while chest muscular endurance was 21.7% higher following HIIT (Fig. 2H: 95% CI = [-11.0, 54.3], *g*=0.79), and seated row muscular endurance was 29.2% higher following HIIT (Fig. 2I: 95% CI = [-13.9, 72.3], *g*=0.91).

#### NK-Cell Numbers and Frequencies

HIIT had a large effect on NK-cell numbers and CD56^dim^ NK-cells. As compared to Controls, the percentage change for the absolute number of CD56+ NK-cells was 51.0% higher following HIIT (Data not shown: 95% CI = [-45.9, 111.0], *g*=0.81), while the frequency of CD56^dim^ NK-cells was 47.9% higher following HIIT (Fig. 3A: 95% CI = [-14.4, 110.0], *g*=0.90), and CD56^bright^ was 36.9% higher following HIIT (Fig. 3A: 95% CI = [-13.9, 61.8], *g*=0.64).

**Figure 3.**
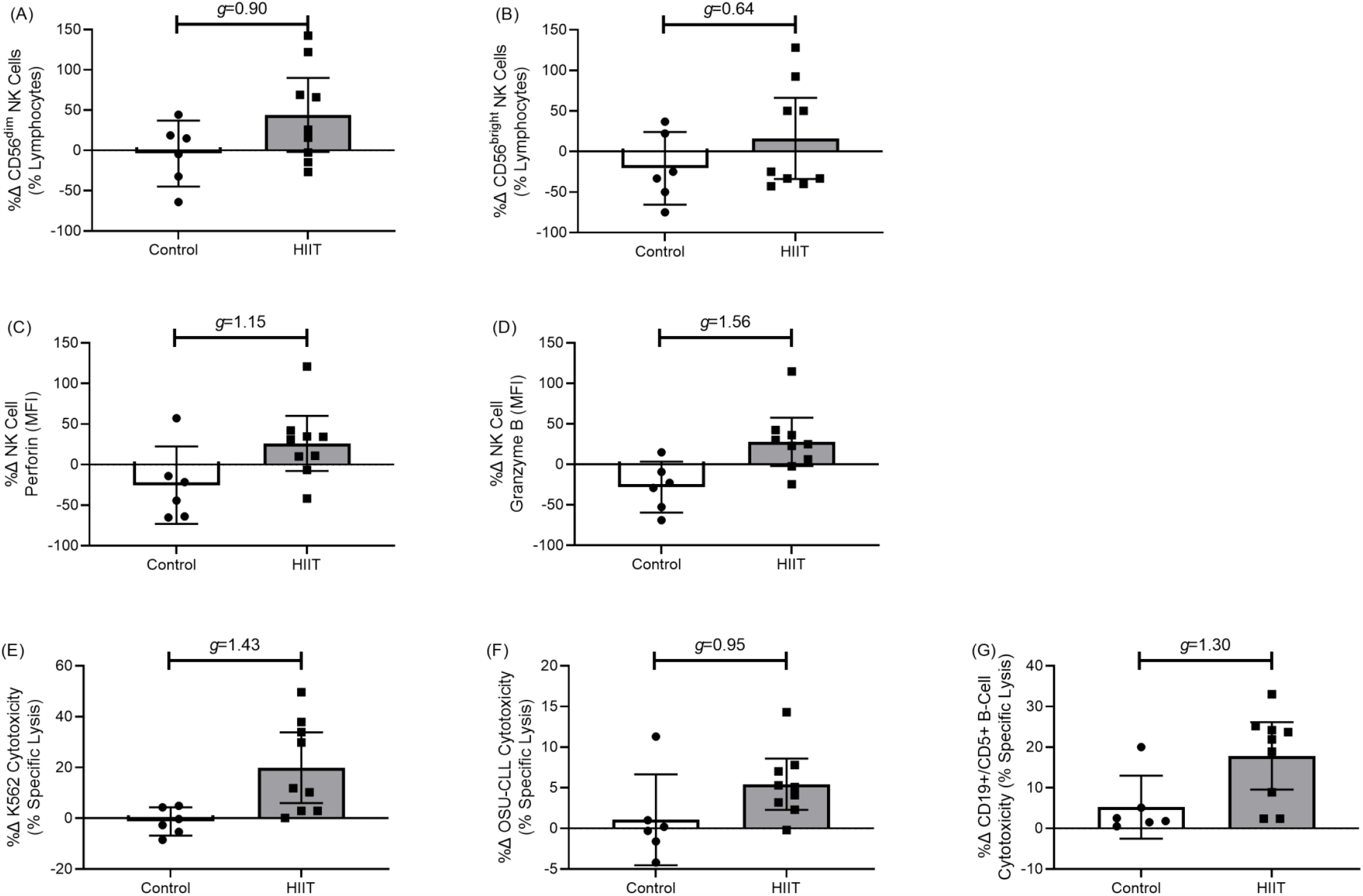
Mean (95% C.I.) percentage change (%Δ) with Hedges G (*g*) group differences between HIIT and Controls for CD56^dim^ NK-cell (A) and CD56^bright^ NK-cell (B) frequencies, expression (MFI) of NK-cell specific perforin (C) and granzyme B (D), and NK-cell cytotoxicity towards K562 (E), OSU-CLL (F), and autologous CD5+ B-cells (G).

#### NK-Cell Functions

HIIT had a large effect on NK-cell functions. As compared to Controls, the percentage change for the expression (MFI) of NK-cell specific perforin was 52.6% higher following HIIT (Fig. 3C: 95% CI = [-3.5, 108.8], *g*=1.15), while expression of granzyme B was 53.8% higher following HIIT (Fig. 3D: 95% CI = [10.1, 97.4], *g*=1.56). As compared to Controls, the percentage change for the NK-cell specific lysis of the K562 cell line was 20.3% higher following HIIT (Fig. 3E: 95% CI = [7.3, 33.3], *g*=1.43), while NK-cell specific lysis of the OSU-CLL cell line was 3.0% higher following HIIT (Fig. 3F: 95% CI = [-1.8, 7.9], *g*=0.95), and NK-cell specific lysis of autologous B-cells was 14.6% higher following HIIT (Fig. 3G: 95% CI = 0.9, 28.4], *g*=1.30).

#### B-cell, T-lymphocyte and Monocyte Changes

HIIT had a small-medium effect on B-cells and T-cells, and a large effect on monocytes. As compared to Controls, the percentage change for the absolute number of CD19+ B-cells was 5.7% lower following HIIT (Fig. 4A: 95% CI = [-38.8, 27.2], *g*=0.21), while the frequency of CD19+ B-cells was 16.9% lower following HIIT (Fig. 4B: 95% CI = [-36.2, 2.4], *g*=0.51), and CD19+/CD5+ CLL-cells was 19.2% lower following HIIT (Fig. 4C: 95% CI = [-40.0, 1.6], *g*=0.54). As compared to Controls, the percentage change for the frequency of CD3+ lymphocytes was 4.4% higher following HIIT (Fig. 4D: 95% CI = [-43.0, 51.8], *g*=0.12), the frequency of CD3+/CD4+ lymphocytes was 10.4% lower following HIIT (Fig. 4E: 95% CI = [-31.7, 10.8], *g*=0.65), the frequency of CD3+/CD8+ lymphocytes was 2.8% higher following HIIT (Fig. 4F: 95% CI = [-36.7, 32.6], *g*=0.26), and the frequency of CD3+/CD56+ lymphocytes was 33.3% higher following HIIT (data not shown: 95% CI = [-37.8, 104.4], *g*=0.66). As compared to Controls, the percentage change for the frequency of CD14+/CD16^neg^ classic monocytes was 7.6% lower following HIIT (Fig. 4G: 95% CI = [-16.7, 1.4], *g*=1.05), while the frequency of CD14+/CD16+ intermediate monocytes was 11.4% higher following HIIT (Fig. 4H: 95% CI = [-21.7, 44.6], *g*=0.89), and the frequency of CD14+/CD16++ non-classic monocytes was 65.5% higher following HIIT (Fig. 4I: 95% CI = [9.2, 121.7], *g*=1.30).

**Figure 4.**
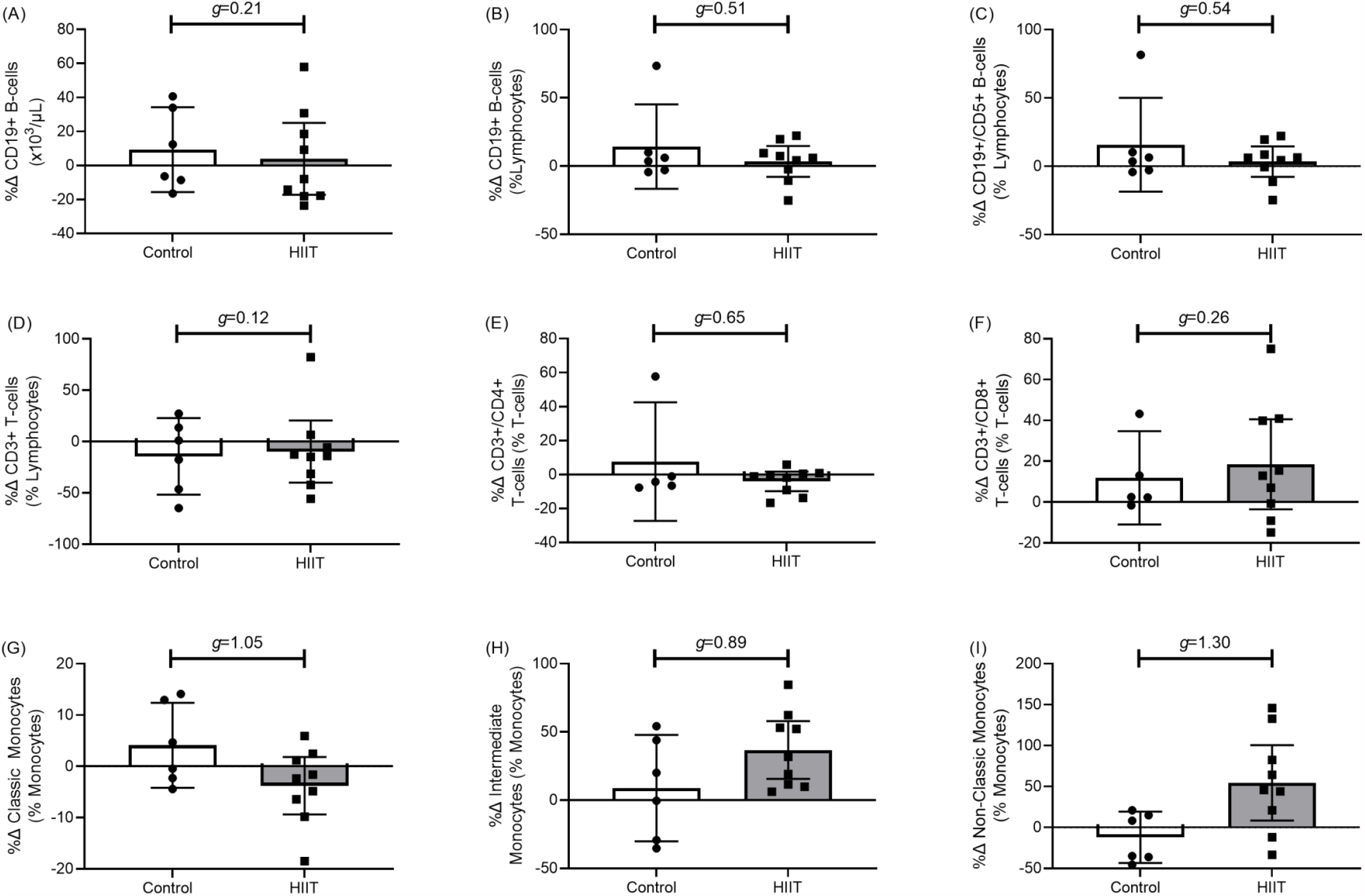
Mean (95% C.I.) percentage change (%Δ) with Hedges G (*g*) group differences between HIIT and Controls for B-cells (A-C), T-cells (D-F), and Monocytes (G-H).

## Discussion

For the first time that we are aware of, we show that 12-weeks of high-intensity interval exercise training (HIIT) combined with muscle endurance-based resistance training is feasible in older adults with treatment naïve CLL. HIIT participants completed 99% of all prescribed exercise sessions/week and minutes/week, at a high-intensity interval heart rate corresponding to 89 (2.8)% of aerobic capacity. We show a high level of compliance for achieving target heart rates that was similar to our previous HIIT studies in non-cancer populations [17, 18, 21]. Our study benefited from a supervised program, with high adherence and retention rates. We adopted a flexible HIIT session scheduling (6am to 6pm, 5 days per week or per request of the participant) and provided supervision by an exercise physiologist. Further, we report the effect sizes of percentage change differences between Controls and HIIT for important physiological and immunological outcomes. As compared to Controls, HIIT had a small-medium effect on aerobic capacity, with higher mean percent changes observed in the Control group. As compared to Controls, HIIT had a large effect on maximal muscle strength and muscular endurance, with higher mean percent changes observed in the HIIT group. Similarly, HIIT had a large effect on monocyte phenotype and NK-cell characteristics, including absolute numbers, tumor cell cytotoxicity and expression of perforin and granzyme B. Taken together, our data support the idea that HIIT is a feasible exercise intervention, and that HIIT is associated with large effects on muscle strength and important immune functions in CLL patients.

### HIIT and Physical Fitness

Patients with cancer typically have low overall fitness that is associated with numerous poor outcomes including fatigue, weight gain and premature mortality [35]. Low fitness is often a consequence of both the cancer and the cancer therapy [35]. Traditional (i.e., low-or moderate-intensity continuous) exercise interventions targeting physical fitness in a range of hematologic malignancies show positive effects for quality of life, physical function, fatigue, cardiovascular fitness, balance, and strength [12-14]. HIIT has recently emerged as an exercise intervention that rapidly increases physical fitness and health outcomes in older adult clinical populations [16]. In cancer populations, the effects of HIIT are less clear. In women undergoing breast cancer therapy [36] or following therapy [37], HIIT did not increase aerobic capacity. However, HIIT increased aerobic capacity for lymphoma and myeloma patients 1-9 months following autologous stem cell transplant [38]. Similar to Persoon and colleagues [38], we observed a small-medium effect for HIIT on aerobic capacity, with both HIIT (5.3%) and Controls (10.3%) increasing. Control group contamination is becoming more common in exercise trials, and here is a likely consequence of the increasing attention for exercise oncology research and the publication of Cancer Physical Activity Guidelines [39]. Given our previous HIIT studies increased aerobic capacity on average 12.4% it is unclear why a similar aerobic exercise program did not elicit similar results [18, 24]. It is possible that the CLL is having a yet unknown effect on cardiorespiratory fitness and future studies should aim to determine the role of CLL on metabolic energy utilization [40]. Unlike our previous HIIT studies, in the current study we included two sessions/week of muscle endurance-building resistance training. As such, HIIT had a large effect on muscular strength and endurance, favoring higher changes in the HIIT group. Muscular strength is an important factor in older adult health and independence, with poor strength associated with increased incidence of frailty and higher risk of premature death [41]. Future studies should aim to determine whether this confers protection from negative outcomes such as frailty, time-to-treatment and premature mortality.

### HIIT and Immune Functions

Although previous exercise interventions in hematologic malignancies show positive effects for aerobic fitness and strength, there is a paucity of data for immune function changes [12-14]. Important immune functions for CLL patients include anti-microbial functions and NK-cell tumor recognition and cytotoxicity [8, 42]. In healthy adults, NK-cells are highly responsive to an acute single bout of exercise that could translate to improvements following chronic exercise training interventions [11]. During and after individual acute bouts of aerobic exercise, NK-cell functions are increased, before returning to pre-exercise levels shortly after exercise cessation [31, 43-46]. For exercise training interventions, effects are less clear. Although some suggest no changes following exercise training [47], others suggest increased NK-cell tumor killing [48]. Similarly, when comparing physically active against physically inactive adults, NK-cell functions can be either higher [49] or similar [50]. In a murine preclinical solid tumor model, Pedersen and colleagues show that 30-days of free-wheel running was associated with up to 60% reduction in tumor burden, mediated by NK-cell improvements [51]. Here, we now demonstrate that HIIT had a large effect on NK-cells, including increased absolute counts, cytotoxic functions, as well as increased perforin and granzyme B expression. Since CLL patients have reduced NK-cell mediated tumor cytotoxicity, reduced expression of activation receptors, and increased inhibitory receptor expression [42, 52], exercise training could be an effective means of reducing primary and/or secondary tumor development.

In addition to NK-cells, we observed that HIIT had a large effect on non-classic (CD14+/CD16++) and classic (CD14+/CD16^neg^) monocytes. Specifically, non-classic monocytes were higher, while classic monocytes were lower following HIIT. These results are the opposite of previous HIIT studies noted in non-cancer participants [18, 21]. The non-classic populations of monocytes are similar to M1 macrophages, and classic monocytes are the primary source of monocyte-derived dendritic cells [53]. In CLL, M1 macrophages are suggested to improve tumor outcomes due to their pro-inflammatory actions in the tumor microenvironment and reducing the balance between M2 macrophages that suppress local immune functions [54]. Future studies should aim to determine the role exercise plays on CLL patient’s monocyte functions, including differentiation, phagocytosis, antigen presentation and cytokine production.

### Strengths and Limitations

Strengths of our study include a focus on treatment naïve CLL patients with a wide range of disease duration (i.e., 6 months to 24 years). As physical dysfunction and poor physical fitness is associated with poor outcomes following commencement of treatment [10], this group of patients reflect a pre-conditioning phase to treatment. As such, improving physical fitness and function may provide better outcomes following treatment. Additionally, our exploratory analyses of immune functions offers a unique insight into how exercise might provide improved resilience to the most common negative outcomes associated with CLL. Limitations of our study include a small number of participants and possible recruitment bias due to non-random assignment of participants to groups. Future randomized trials are warranted to demonstrate the effects of HIIT on physical fitness and immune function as they relate to physical resilience in CLL. Specifically, future incidence of infections, secondary malignancies, and time-to-treatment.

### Conclusion

In conclusion, we demonstrate that 12-weeks of HIIT combined with muscle endurance-based resistance training for treatment naïve CLL patients is feasible, and is associated with large effects on muscle strength and normal immune cell functions. Our findings require confirmation in an adequately powered randomized trial to determine the benefits of HIIT on important physical and immunological outcomes in CLL.

## Data Availability

Data will be made available upon reasonable request to the study team

## List of Abbreviations

B2M: β2-microglobulin
CD: cluster of differentiation
CLL: Chronic Lymphocytic Leukemia
CLL-IPI: Chronic Lymphocytic Leukemia – International Prognostic Index
CPET: cardiopulmonary exercise test
DMSO: Dimethyl sulfoxide
DPBS: Dulbecco’s phosphate buffered saline
EBV: Epstein Barr Virus
EDTA: Ethylenediaminetetraacetic acid
ER: Emergency Room
FBS: fetal bovine serum
FMO: flow minus one
HIIT: high intensity interval training
ICAM-1: Intercellular Adhesion Molecule 1
IGHV: immunoglobulin heavy chain gene
IL: interleukin
NK-cell: Natural Killer Cell
PBS: phosphate buffered saline
OSU-CLL: Ohio State University-Chronic Lymphocytic Leukemia
PBMC: peripheral blood mononuclear cell
RPMI: Roswell Park Memorial Institute Medium
VO2peak: estimated peak volume of oxygen consumption

## Declarations

### Ethics Statement

The study involving human participants was reviewed and approved by the Duke University Medical Center Institutional Review Board. The patients/participants provided their written informed consent prior to participating in this study.

### Consent for publication

Not applicable

## Availability of Data and Materials

The datasets generated during the present study are not publicly available, owing to the risk of disclosure or deduction of private individual information, but they are available from the corresponding author on reasonable request.

## Authors’ Contributions

DBB, DMB and AS conceived and designed the study and experimental approach. GM, MD, and SF performed the physiological and functional testing and exercise training of participants. GM managed participant scheduling and data collection. AS, JBW, and DMB provided clinical guidance and access to their patients. DBB and EH performed the immunological experiments. CP assisted in the statistical analyses. All authors contributed to critical revisions and approval of the final manuscript.

## Acknowledgments

We acknowledge the staff members at the Duke Center for Living and the Duke Cancer Institute for their help with patient care. We appreciate the support of the Department of Medicine at Duke University and, of course, all the participants.

## References

1. Howlader N, et al. SEER Cancer Statistics Review, 1975-2017, National Cancer Institute. Bethesda, MD. 2020 [cited 2020; Available from: https://seer.cancer.gov/csr/1975_2017/.

2. Scarfò, L., A.J. Ferreri, and P. Ghia, Chronic lymphocytic leukaemia. Crit Rev Oncol Hematol, 2016. 104: p. 169–82.

3. Washburn, L., Chronic lymphocytic leukemia: the most common leukemia in adults. Jaapa, 2011. 24(5): p. 54–8 Quiz 59.

4. Shanafelt, T.D., et al., Age at diagnosis and the utility of prognostic testing in patients with chronic lymphocytic leukemia. Cancer, 2010. 116(20): p. 4777–87.

5. Dighiero, G., et al., Chlorambucil in indolent chronic lymphocytic leukemia. French Cooperative Group on Chronic Lymphocytic Leukemia. N Engl J Med, 1998. 338(21): p. 1506–14.

6. Shustik, C., et al., Treatment of early chronic lymphocytic leukemia: intermittent chlorambucil versus observation. Hematol Oncol, 1988. 6(1): p. 7–12.

7. Solomon, B.M., et al., Overall and cancer-specific survival of patients with breast, colon, kidney, and lung cancers with and without chronic lymphocytic leukemia: a SEER population-based study. J Clin Oncol, 2013. 31(7): p. 930–7.

8. Rossi, D., et al., Early stage chronic lymphocytic leukaemia carrying unmutated IGHV genes is at risk of recurrent infections during watch and wait. Br J Haematol, 2008. 141(5): p. 734–6.

9. Riches, J.C. and J.G. Gribben, Immunomodulation and immune reconstitution in chronic lymphocytic leukemia. Semin Hematol, 2014. 51(3): p. 228–34.

10. Goede, V., et al., Evaluation of geriatric assessment in patients with chronic lymphocytic leukemia: Results of the CLL9 trial of the German CLL study group. Leuk Lymphoma, 2016. 57(4): p. 789–96.

11. Duggal, N.A., et al., Can physical activity ameliorate immunosenescence and thereby reduce age-related multi-morbidity? Nat Rev Immunol, 2019.

12. Sitlinger, A., D.M. Brander, and D.B. Bartlett, Impact of Exercise on the Immune System and Outcomes in Hematologic Malignancies. Blood Advances, 2020. 4(8): p. 1801–1811.

13. Streckmann, F., et al., Exercise program improves therapy-related side-effects and quality of life in lymphoma patients undergoing therapy. Ann Oncol, 2014. 25(2): p. 493–9.

14. Courneya, K.S., et al., Randomized controlled trial of the effects of aerobic exercise on physical functioning and quality of life in lymphoma patients. J Clin Oncol, 2009. 27(27): p. 4605–12.

15. Gleeson, M., et al., The anti-inflammatory effects of exercise: mechanisms and implications for the prevention and treatment of disease. Nat Rev Immunol, 2011. 11(9): p. 607–615.

16. Campbell, W.W., et al., High-Intensity Interval Training for Cardiometabolic Disease Prevention. Med Sci Sports Exerc, 2019. 51(6): p. 1220–1226.

17. Bartlett, D.B., et al., Rejuvenation of Neutrophil Functions in Association with Reduced Diabetes Risk Following Ten Weeks of Low-Volume High Intensity Interval Walking in Older Adults with Prediabetes – A Pilot Study. Frontiers in Immunology, 2020. 11(729).

18. Bartlett, D.B., et al., Ten weeks of high-intensity interval walk training is associated with reduced disease activity and improved innate immune function in older adults with rheumatoid arthritis: a pilot study. Arthritis Res Ther, 2018. 20(1): p. 127.

19. Robinson, E., et al., Short-term high-intensity interval and moderate-intensity continuous training reduce leukocyte TLR4 in inactive adults at elevated risk of type 2 diabetes. J Appl Physiol (1985), 2015. 119(5): p. 508–16.

20. Bouaziz, W., et al., Effect of high-intensity interval training and continuous endurance training on peak oxygen uptake among seniors aged 65 or older: A meta-analysis of randomized controlled trials. Int J Clin Pract, 2020: p. e13490.

21. Bartlett, D.B., et al., Neutrophil and Monocyte Bactericidal Responses to 10-Weeks of Low-Volume High Intensity Interval or Moderate-Intensity Continuous Training in Sedentary Adults. Oxid Med Cell Longev, 2017. 2017: p. 12.

22. Ralston, G.W., et al., Weekly Training Frequency Effects on Strength Gain: A Meta-Analysis. Sports medicine - open, 2018. 4(1): p. 36–36.

23. Hallek, M., et al., Guidelines for diagnosis, indications for treatment, response assessment and supportive management of chronic lymphocytic leukemia. Blood, 2018.

24. Bartlett, D.B., et al., Rejuvenation of Neutrophil Functions in Association With Reduced Diabetes Risk Following Ten Weeks of Low-Volume High Intensity Interval Walking in Older Adults With Prediabetes – A Pilot Study. Frontiers in Immunology, 2020. 11(729).

25. Pescatello, L.S. and M. American College of Sports, ACSM’s guidelines for exercise testing and prescription. 2014, Philadelphia: Wolters Kluwer/Lippincott Williams & Wilkins Health.

26. Knutzen, K.M., L.R. Brilla, and D. Caine, Validity of 1RM Prediction Equations for Older Adults. The Journal of Strength & Conditioning Research, 1999. 13(3): p. 242–246.

27. Howley, E.T., Type of activity: resistance, aerobic and leisure versus occupational physical activity. Med Sci Sports Exerc, 2001. 33(6 Suppl): p. S364-9; discussion S419-20.

28. Bartlett, D.B. and N.A. Duggal, Moderate physical acticity is associated with increased naive: memory T-cell ratio in healthy old; potential role of IL-15. Age and Ageing, 2020. 49(3): p. 368–373.

29. Hazeldine, J., P. Hampson, and J.M. Lord, Reduced release and binding of perforin at the immunological synapse underlies the age-related decline in Natural Killer cell cytotoxicity. Aging Cell, 2012.

30. Hertlein, E., et al., Characterization of a New Chronic Lymphocytic Leukemia Cell Line for Mechanistic In Vitro and In Vivo Studies Relevant to Disease. PLOS ONE, 2013. 8(10): p. e76607.

31. Bigley, A.B., et al., Acute exercise preferentially redeploys NK-cells with a highly-differentiated phenotype and augments cytotoxicity against lymphoma and multiple myeloma target cells. Brain Behav Immun, 2014. 39: p. 160–71.

32. group, I.C.-I.w., An international prognostic index for patients with chronic lymphocytic leukaemia (CLL-IPI): a meta-analysis of individual patient data. Lancet Oncol, 2016. 17(6): p. 779–790.

33. Hedges, L.V., Distribution Theory for Glass’s Estimator of Effect Size and Related Estimators. Journal of Educational Statistics, 1981. 6(2): p. 107–128.

34. Durlak, J., How to select, calculate, and interpret effect sizes. Journal of pediatric psychology, 2009. 34 9: p. 917–28.

35. Schmitz, K.H., Exercise Oncology: Prescribing Physical Activity Before and After a Cancer Diagnosis. 2020: Springer International Publishing.

36. Lee, K., et al., Feasibility of high intensity interval training in patients with breast Cancer undergoing anthracycline chemotherapy: a randomized pilot trial. BMC Cancer, 2019. 19(1): p. 653.

37. Schmitt, J., et al., A 3-week multimodal intervention involving high-intensity interval training in female cancer survivors: a randomized controlled trial. Physiol Rep, 2016. 4(3).

38. Persoon, S., et al., Randomized controlled trial on the effects of a supervised high intensity exercise program in patients with a hematologic malignancy treated with autologous stem cell transplantation: Results from the EXIST study. PLOS ONE, 2017. 12(7): p. e0181313.

39. Campbell, K.L., et al., Exercise Guidelines for Cancer Survivors: Consensus Statement from International Multidisciplinary Roundtable. Med Sci Sports Exerc, 2019. 51(11): p. 2375–2390.

40. Sitlinger, A., et al., Physiological Fitness and the Pathophysiology of Chronic Lymphocytic Leukemia (CLL). Cells, 2021. 10(5): p. 1165.

41. Li, R., et al., Associations of Muscle Mass and Strength with All-Cause Mortality among US Older Adults. Medicine and science in sports and exercise, 2018. 50(3): p. 458–467.

42. Huergo-Zapico, L., et al., Expansion of NK cells and reduction of NKG2D expression in chronic lymphocytic leukemia. Correlation with progressive disease. PLoS One, 2014. 9(10): p. e108326.

43. Gleeson, M. and N.C. Bishop, The T cell and NK cell immune response to exercise. Ann Transplant, 2005. 10(4): p. 43–8.

44. Evans, E.S., et al., Impact of Acute Intermittent Exercise on Natural Killer Cells in Breast Cancer Survivors. Integr Cancer Ther, 2015. 14(5): p. 436–45.

45. Shephard, R.J. and P.N. Shek, Effects of exercise and training on natural killer cell counts and cytolytic activity: a meta-analysis. Sports Med, 1999. 28(3): p. 177–95.

46. Nieman, D.C., et al., Effects of high-vs moderate-intensity exercise on natural killer cell activity. Med Sci Sports Exerc, 1993. 25(10): p. 1126–34.

47. Campbell, P.T., et al., Effect of exercise on in vitro immune function: a 12-month randomized, controlled trial among postmenopausal women. J Appl Physiol (1985), 2008. 104(6): p. 1648–55.

48. Woods, J.A., et al., Effects of 6 months of moderate aerobic exercise training on immune function in the elderly. Mech Ageing Dev, 1999. 109(1): p. 1–19.

49. Moro-Garcia, M.A., et al., Frequent participation in high volume exercise throughout life is associated with a more differentiated adaptive immune response. Brain Behav Immun, 2014. 39: p. 61–74.

50. Yan, H., et al., Effect of moderate exercise on immune senescence in men. Eur J Appl Physiol, 2001. 86(2): p. 105–11.

51. Pedersen, L., et al., Voluntary Running Suppresses Tumor Growth through Epinephrine-and IL-6-Dependent NK Cell Mobilization and Redistribution. Cell Metabolism, 2016. 23(3): p. 554–562.

52. Wensveen, F.M., V. Jelenčić, and B. Polić, NKG2D: A Master Regulator of Immune Cell Responsiveness. Frontiers in Immunology, 2018. 9(441).

53. Boyette, L.B., et al., Phenotype, function, and differentiation potential of human monocyte subsets. PloS one, 2017. 12(4): p. e0176460–e0176460.

54. Petty, A.J. and Y. Yang, Tumor-Associated Macrophages in Hematologic Malignancies: New Insights and Targeted Therapies. Cells, 2019. 8(12): p. 1526.

